# Modelling Disease progression of Multiple Sclerosis in a South Wales Cohort

**DOI:** 10.1101/2023.09.15.23295414

**Authors:** Emeka C Uzochukwu, Katharine E Harding, James Hrastelj, Karim L Kreft, Peter Holmans, Lesley Jones, Neil P Robertson, Emma C Tallantyre, Michael Lawton

## Abstract

**Objectives:** To model multiple sclerosis (MS) disease progression and compare disease trajectories by sex, age of onset, and year of diagnosis.

**Study Design and settings:** Longitudinal EDSS scores were collected since 1985 for relapse-onset MS patients at MS clinics in South Wales and modelled using a multilevel model (MLM). The MLM adjusted for baseline covariates (sex, age of onset, year of diagnosis, and disease modifying treatments (DMTs)), and included interactions between baseline covariates and time variables.

**Results:** The optimal model was truncated at 30 years after disease onset and excluded EDSS recorded within 3 months of relapse. As expected, older age of onset was associated with faster disease progression at 15 years (effect size (ES): 0.75; CI: 0.63, 0.86; P: <0.001) and female sex progressed more slowly at 15 years (ES: -0.43; CI: -0.68, -0.18; P: <0.001). Patients diagnosed more recently (defined as 2007-2011 and >2011) progressed more slowly than those diagnosed historically (<2006); (ES: -0.46; CI: -0.75, -0.16; P: 0.006) and (ES: -0.95; CI: -1.20, -0.70; P: <0.001), respectively.

**Conclusion:** We present a novel model of MS outcomes, accounting for the nonlinear trajectory of MS and effects of baseline covariates, validating well-known risk factors (sex and age of onset) associated with disease progression. Also, patients diagnosed more recently progressed more slowly than those diagnosed historically.

## Introduction

Multiple sclerosis (MS), a chronic inflammatory disorder affecting the central nervous system (CNS), is one of the leading cause of neurological disability in young adults [1]. Its aetiology remains unclear, but both genetic and environmental factors are contributors. Onset is most frequent in the third and fourth decades of life. Females are almost three times more frequently affected by MS. Pathologically, MS is characterised by focal areas of inflammation and demyelination as well as more diffuse neurodegenerative features, throughout the CNS. Early MS commonly presents as recurrent sub-acute episodes of neurological dysfunction, which may remit to a varying degree [2]. Most people with MS (PwMS) develop a secondary progressive phase after a variable interval. A significant minority of patients present with primary progressive MS (PPMS) [3]. Moreover, the presentation and subsequent disease course of MS are highly variable and difficult to predict at onset, making decisions on management challenging for clinicians and PwMS [4].

Since the emergence of disease modifying therapies (DMTs) for MS in the 1990’s, an increasing number of DMTs with variable efficacy and safety profile are now available [5], [6]. However, uncertainty remains over selection and sequencing of DMTs; aggressive immune suppression early in MS may affect longer-term disease outcomes, but some high-efficacy DMTs have risks of adverse events [7], [8]. There is an urgent need to identify biomarkers that can inform disease prognosis, treatment response and potential for adverse events to enable a more personalised approach to interventions.

Reliable and contemporary data on MS outcomes is needed for patient counselling, disease management, exploring the utility of candidate biomarkers, and to establish real-world effectiveness of new treatment or interventions. The expanded disability status scale (EDSS [9]) remains an internationally recognised clinical disablility outcome measure in MS and a key element of epidemiological studies, clinical trial design and treatment approval [10], despite having relatively poor intra- and inter-rater reliability, over-reliance on mobility, and low sensitivity to vision, arm function, or cognition [11].

In previous multilevel modelling (MLM) studies, we were able to highlight the nonlinear trajectory of EDSS scores in PwMS eligible for DMTs and demonstrate modest benefits for injectable DMTs [12], [13]. In the current prospective cohort study of both treated and untreated PwMS, we aimed to develop a MLM using contemporary MS data incorporating additional demographic data (sex, age of onset, and year of diagnosis) to investigate differences in disease trajectory.

## Methods

### Data source/measurements

Data have been collected as part of an ongoing prospective study since 1985, [14]. Patients are reviewed annually from diagnosis and data on demographics, relapses, disease course and disability were collected at each encounter. EDSS is routinely assessed and recorded in a standardised web-based form and incorporated into the registry. Inclusion criteria in this study were: a diagnosis of MS, a recorded date of MS onset, and consented for data to be used for research. Individuals were followed up either to October 2021, death, loss to follow-up, or withdrawal from the study. Individuals with PPMS were excluded as this minority progress more rapidly compared to relapse-onset MS (ROMS) patients [15]. This study has been approved by the Research Ethics Committee (REC approval number: 19/WA0289). Time since onset (years) was used as the time-metric. DMTs were time-varying covariate because individuals were administered DMTs at varying time points. Other baseline covariates included sex, age of onset, and year of diagnosis. Time since onset and age of onset were continuous covariates, while sex and year of diagnosis (1986 – 2006, 2007 - 2011, >2011) were categorical covariates. The boundaries of year of diagnosis were based on the publication of revised diagnostic criteria[16], [17]. In general, with updated diagnostic criteria, shorter time between onset, diagnosis and initiation of treatment was achieved, which may affect long-term disability [18].

## Development of the statistical model

### Statistical analysis

Association between EDSS disability outcome and covariates was assessed using MLMs, which are widely used for modelling disease progression [19], [20]. Several MLMs were developed, and interactions were tested (Supplementary Table 1). To handle relapses, EDSS scores recorded within 1-, 3-, and 6-months post-relapse were removed in a series of MLMs to ascertain the optimal window.

Autocorrelation was handled using median EDSS within quarter-year intervals. The best fitting model was identified using Akaike Information Criteria (AIC), root mean square error (RMSE), proportion of EDSS scores within ±0.5 predicted EDSS (PWPE) and the proportion outside ±2 predicted EDSS (POPE). Nonconstant within-person variance was accounted for using a complex level 1 variance model (CLOVM). Statistical analyses and model fitting were performed using R version 4.1.1 [21], R2MLwiN [22] and lme4 [23].

### Multilevel model

MLM is a common statistical model for handling dependent observations, it can summarise the trajectory of the response over time, and account for within and between individual variability, thereby providing better estimates and predictions compared to the classical linear regression model [26]. Since EDSS scores vary regarding presentation and disease trajectories, we initially assumed a random intercept and slope model with time of measurement at level 1 and individuals at level 2:

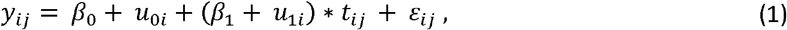

where *y*_*ij*_ denotes the EDSS score at time *j* for individual *i*, and *t*_*ij*_ the time for individual *i* at time *j*. The *β*_0_ is the overall intercept, and *β*_1_ is the overall slope. The *u*_0*i*_ and *u*_1*i*_ are individual-specific random effects assumed to follow a bivariate normal distribution:

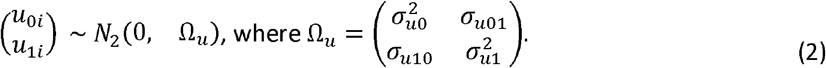

Thus, *β*_0_ + *u*_0*i*_ and *β*_1_ + *u*_1*i*_ are the intercept and slope for individual *i. ε*_*ij*_, the observation-level residual, is assumed to follow a normal distribution with mean zero and variance 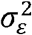.

### Time metric and transformations

Time since onset was used as the time variable and due to differences in disease trajectory for pwMS, fractional polynomials were allowed to model nonlinear relationships between EDSS scores and time, and a value of 1 was added to the time variable to handle zeros in the data [27]. The fractional polynomial approach was preferred to splines since it is a simple way to choose among different transformations, produces smooth monotonic curves and avoids selecting location and knots necessary in splines [28].

### Relapses and Autocorrelation structure

Stable recovery from relapse for most patients is achieved within 3 months [32], although some continue to recover beyond 6 months [34]. To avoid modelling EDSS in relapse all scores within 1-, 3-, and 6-months post-relapse were omitted, and the optimal window was selected according to the magnitude of the intercept-variance of the observations.

The observations could be taken at short intervals or remain unchanged for long periods causing autocorrelation, which could be accounted for using autoregressive-moving average models [33] or an integrated Ornstein-Uhlenbeck stochastic process [34]. We reduce autocorrelation by summarising observations within quarter-year intervals using the median score for individuals before models fitting.

### Complex level 1 variation

The variation within individuals may depend on other covariates such as time, which allows nonconstant variance in residuals [35]. We accounted for nonconstant variance for the time of measurement using the CLOVM, which models level 1 residuals as a function of time [36], and used fractional polynomials to determine the optimal time function. Complex level 1 variation (CLOV) can be caused by measurement error and changes in EDSS scores over time, as disability level ranges considerably [18] and misclassification may be present with more variation than expected at the lower end of the EDSS scale versus the higher end due to intra- and inter-rater differences [11].

### Effect of patient characteristics on progression

When trajectories are non-linear over time with more than one time term it becomes more difficult to look at the effect a variable (such as sex) has on progression because that variable can have an effect on multiple terms in the model (e.g intercept, square root of time and log of time). So, when considering the effect of sex, age of onset and year of diagnosis on progression we have considered that effect at onset and at a time of 15 years post onset. The effect at a time of 15 years post onset on EDSS is a combination of the effect on the intercept and any interactions with time.

## Results

A total of 2,293 PwMS were identified with consent to research, of whom 2,167 had a recorded date of onset. After filtering, a total of 20836 EDSS scores from 1,787 PwMS were included in the analysis.

Patient demographics are summarised in Table 1. A total of 1,787 patients with 20,836 EDSS scores from 1985 to 2021 were included. The mean age of onset was 31.5 years, and the majority were females (71.9%). Median follow-up time was 7.6 years, with 58.7% and 36.9% followed-up for over 5 and 10 years, respectively. DMTs were administered to 35.1% of the whole cohort since 1985.

**Table 1:**
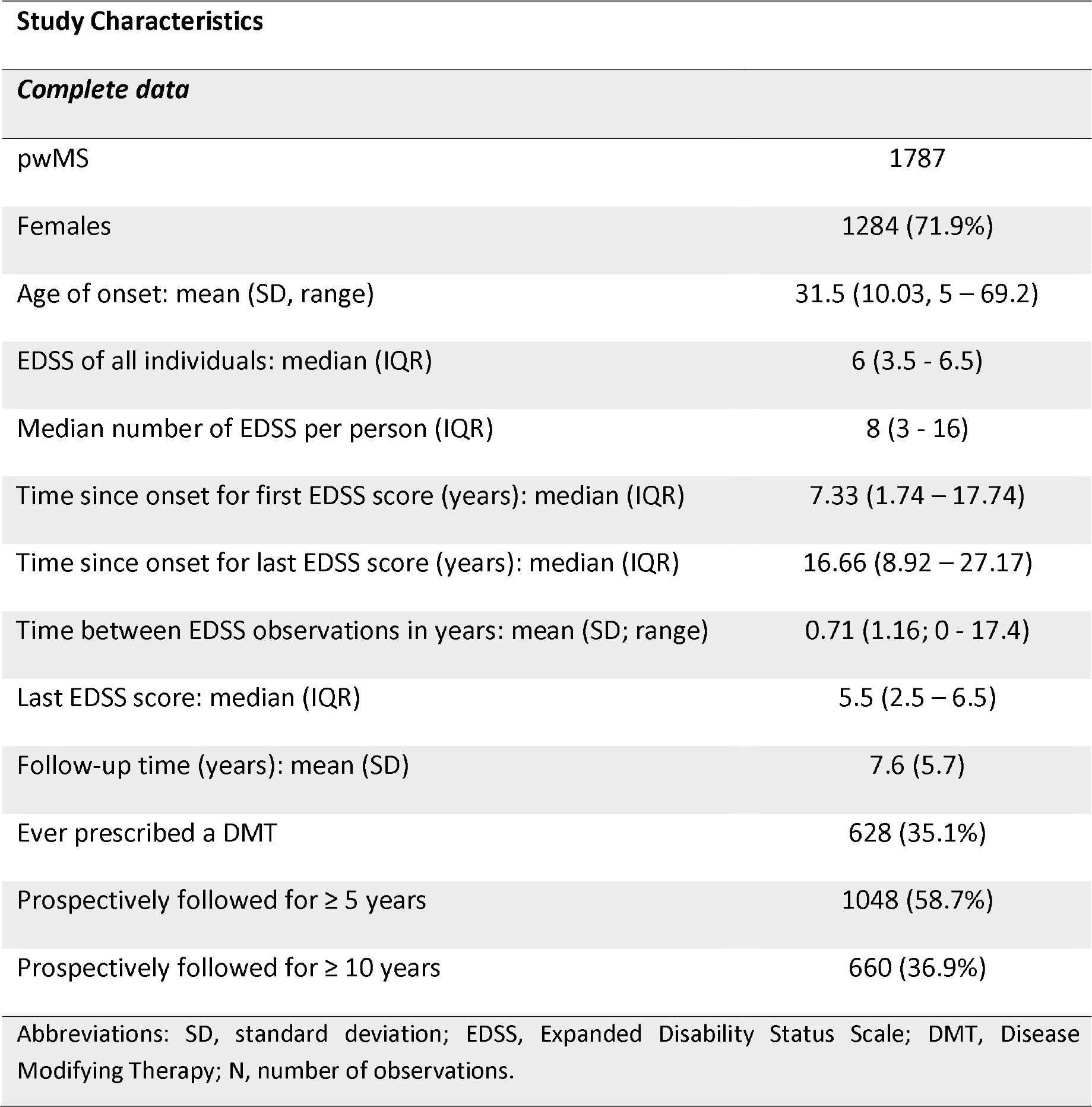
Summary statistics of patient demographics.

To model progression while ignoring relapses, EDSS scores recorded within 1-month post-relapse were excluded, reducing the number EDSS scores to 18,852. To reduce autocorrelation, EDSS scores within quarter-year intervals were summarised using the median, reducing the number of EDSS scores to 15,817 (Figure 1).

**Figure 1:**
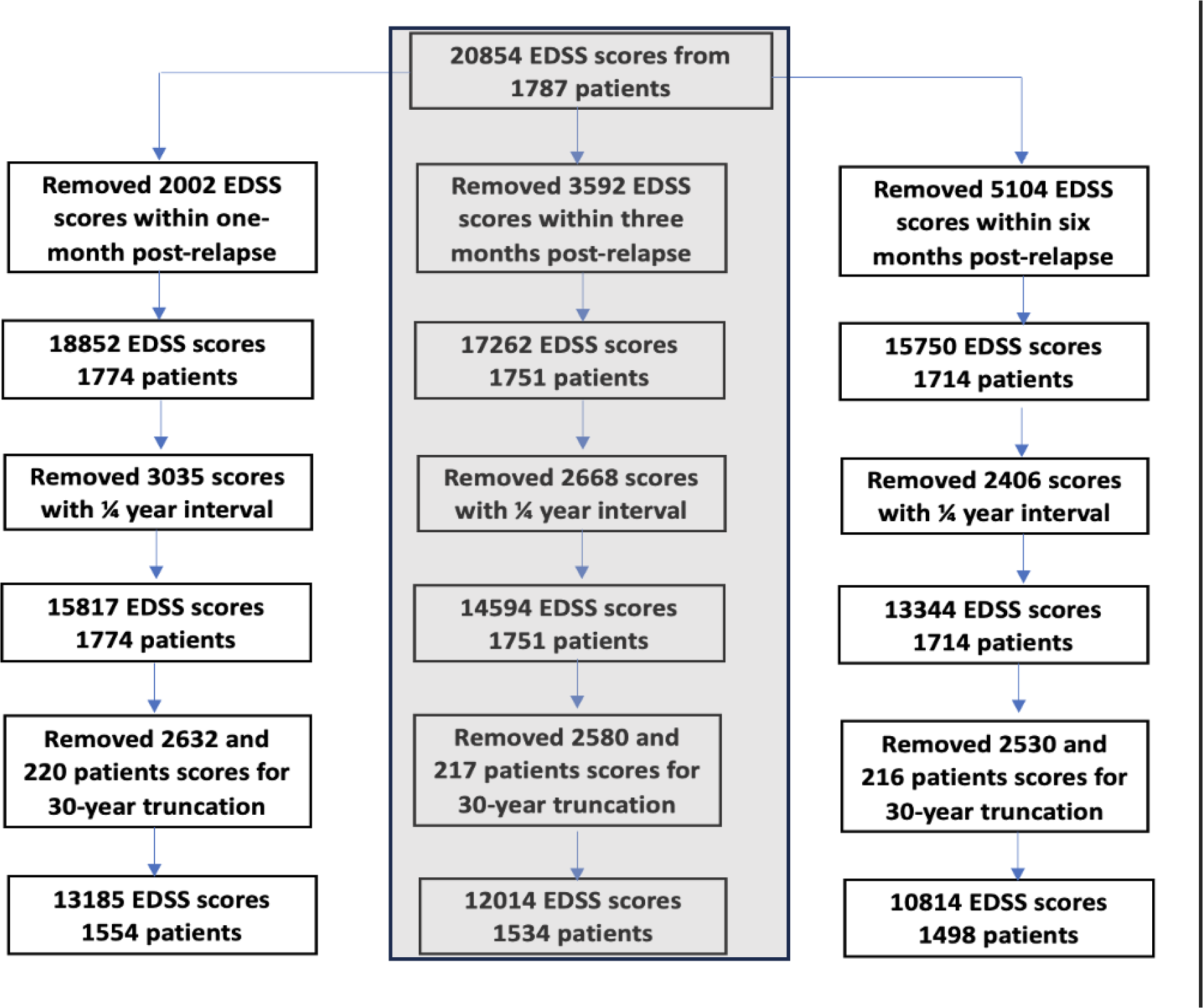
Flow chart of the different models compared within this study (second column was used in the final model).

The CLOV MLM with square root and log of time transformations with interactions was the best model; with better fitted values compared to linear fixed effect model (Supplementary Table 1 and Figure 1). However, accounting for CLOV in models with nonlinear time proved difficult due to outlying EDSS observations in time. Therefore, observations recorded beyond 30 years for time since onset were excluded in subsequent analyses (Supplementary Table 2 and Figure 2).

Because effects of relapse could continue beyond 1-month, EDSS scores within 3- and 6-months post-relapse were removed, and the model refitted. The 3-months post-relapse model was chosen as the optimal model because the CI of the intercept-variance at level 1 for 3- and 6-months are similar (CI: 0.76, 0.89) and (CI: 0.77, 0.84), respectively (Supplementary Table 4). Individuals on DMT showed faster progression than those not exposed to DMT, however, the effect size is small, and the P-value is only borderline significant (ES: 0.06; P: 0.04, CI: 0.001, 0.10). PwMS who never had a DMT were followed up less frequently and had fewer EDSS measurements; their rate of EDSS measurements was ES: 0.57 (P: <0.001; CI: 0.55, 0.58) times that of patients who have had DMT.

The combined effects of covariates and their interactions were tested jointly (Table 2). Sex and sex-time interactions suggested no significant difference between sexes at onset, although, at 15 years females were associated with lower EDSS progression. Similarly, the combined effect of age of onset at 15 years indicate that of age of onset influences progression. At onset, the combined effect of year of diagnosis were not significantly different for patients diagnosed historically (1986-2006) compared to recently diagnosed patients (2007-2011, 2012-2018, and >2018), yet patients diagnosed historically progressed faster than patients diagnosed under the (revised) McDonald criteria. Comparing EDSS scores for patients diagnosed after 2018 to diagnosis made between 2007-2011 and 2012-2018 showed no significant difference at the initial year. However, at 10 years post-onset diagnosis made after 2018 had progressed more slowly compared to 2007-2011, but not 2012-2018 (P: 0.11; CI: -0.72, 0.22). Hence, we combined patients diagnosed >2018 with those diagnosed 2012-2018 in further analysis. The crude effects of these three covariates, when not adjusted for each other or DMT use, were very similar (Supplementary Table 5).

**Table 2:** Combined effects of covariates (P-value & 95% CI) of complex level 1 variance models with *Var*(*t*) = 0 using 30-year truncated data.

| Variables | Effect estimate (95% CI) | P-value |
| --- | --- | --- |
| Female (onset) | 0.47 (-0.10, 1.0) | 0.11 |
| Female (year 15) | -0.43 (-0.68, -0.18) | 0.001 |
| Age of onset, one SD increase (onset) | 0.07 (-0.19, 0.33) | 0.60 |
| Age of onset, one SD increase (year 15) | 0.73 (0.62, 0.85) | <0.001 |
| **Comparing 1986 – 2006 to other year-categories** |  |  |
| Year of diagnosis (2007 – 2011) (onset) | -0.64 (-1.41, 0.14) | 0.11 |
| Year of diagnosis (>2011) (onset) | -0.40 (-1.04, 0.25) | 0.23 |
| Year of diagnosis (2007 – 2011) (year 10) | -0.45 (-0.75, -0.14) | 0.004 |
| Year of diagnosis (>2011) (year 10) | -0.90 (-1.15, -0.65) | <0.001 |

We compared the effect of variables on EDSS at onset and 15 years post onset for sex and age of onset, 10 years post onset for year of diagnosis, using their associations with intercept and time interactions.
Abbreviations: CI, Confidence interval; SD, Standard Deviation

The average trajectories of both sexes (Figure 2a) indicate that females have higher EDSS scores at onset but progressed more slowly with average EDSS scores of 5.1 and 4.8 for males and females, respectively, at 15 years. Furthermore, individuals with age of onset above the average had higher EDSS at 15 years post onset compared to patients with an age of onset below the average, with average EDSS scores of approximately 3.9 and 5.7, respectively (Figure 2b). A comparison of the average trajectories for the categorised year of diagnosis showed a faster progression for patients diagnosed historically compared to patients diagnosed more recently (2007-2011 and >2011), with average EDSS scores of approximately 4.0, 3.5, and 3.0, respectively, 10 years post onset (Figure 2c).

**Figure 2a:**
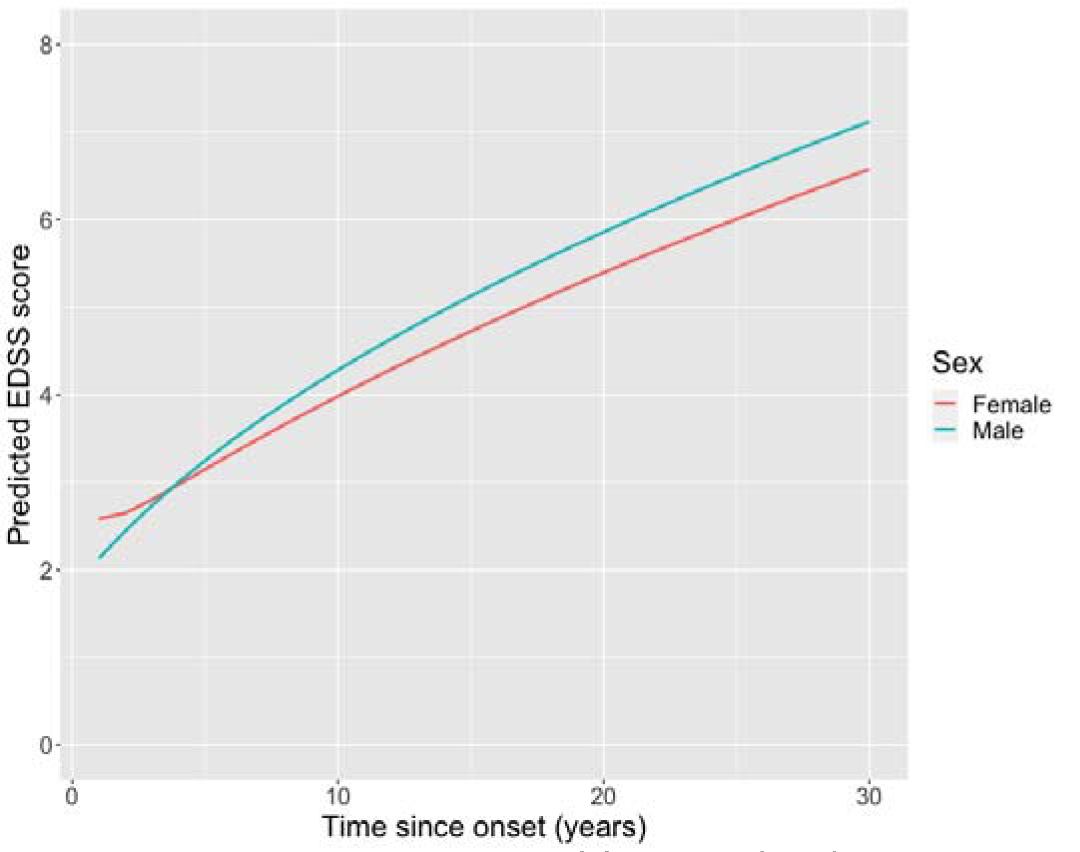
Progression plot of females (red) and males (green)

**Figure 2b:**
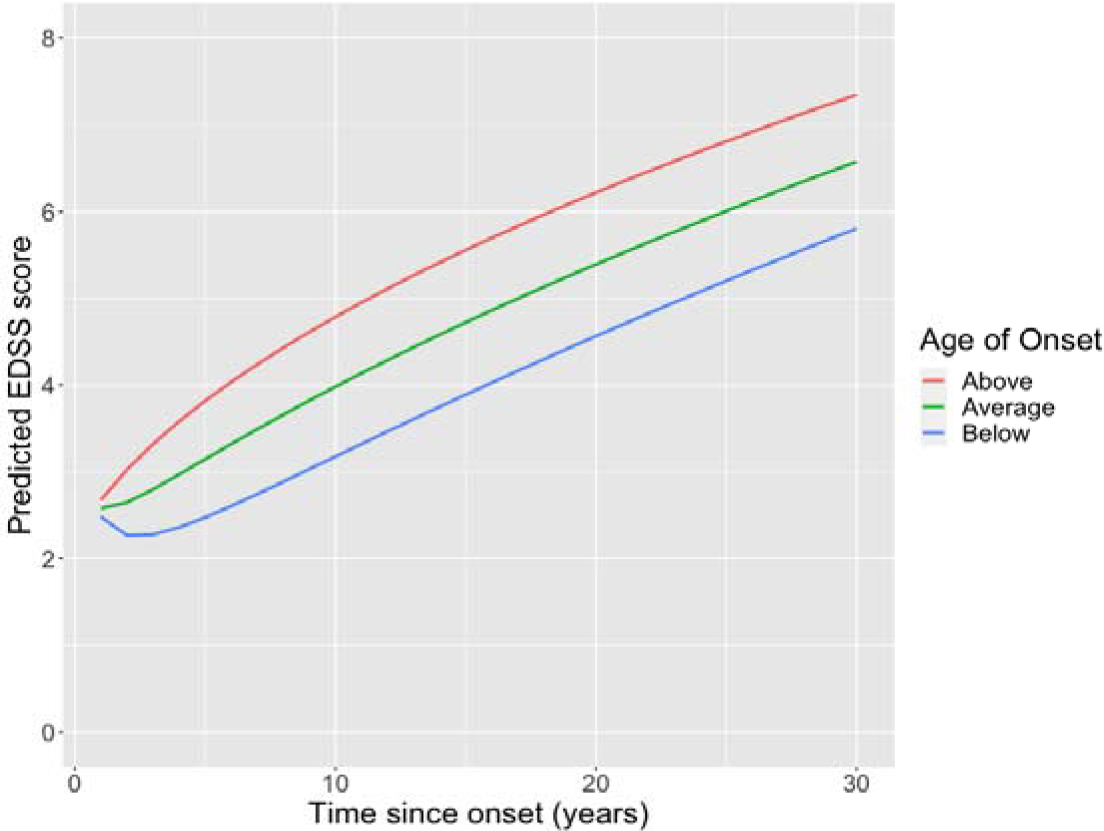
Progression plot of patients young (blue) and older (red) at onset.

**Figure 2c:**
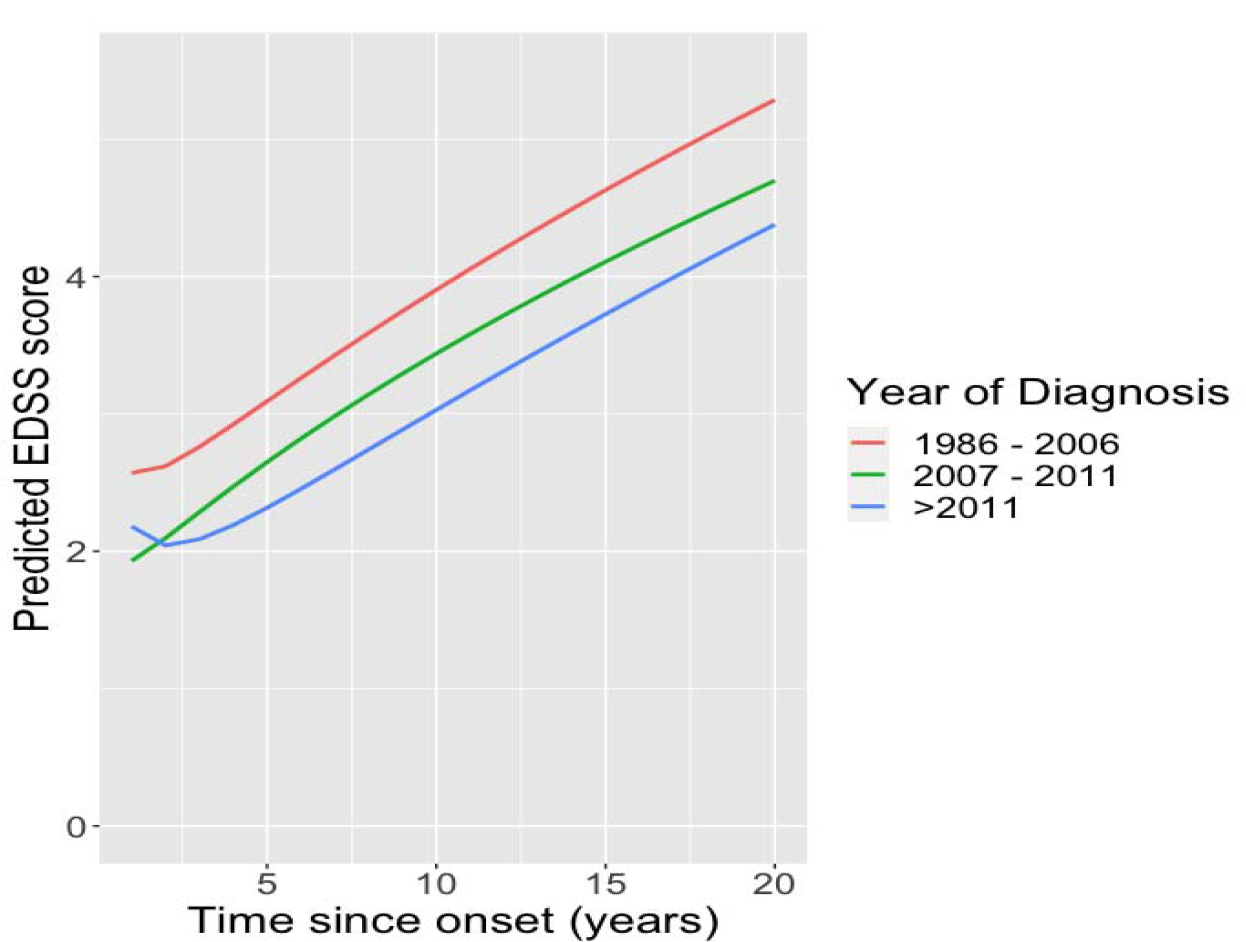
Progression plot for year of diagnosis. 1986 - 2006 (red), 2007 - 2011 (green), >2011 (blue).

## Discussion

The purpose of this study was to develop a model MS for disease progression in a contemporary population-based cohort and assess the differences in disability trajectory using baseline covariate (sex, age of onset and year of diagnosis). Overall, we show that patients diagnosed historically appear to have progressed faster compared to those diagnosed more recently and validated previous studies that females progressed more slowly than males, and patients with above average age of onset reached disability milestones faster than patients with below average age of onset.

Time since onset (years) was the time variable and fractional polynomials were used to allow flexible trajectories of progression in individuals. Models were developed after removing observations within 1-, 3-, and 6-months post-relapse to select the optimal recovery window. The 3-months relapse window was chosen because the CI of the intercept variance at level 1 overlaps with that of 6-months relapse window. Moreover, earlier findings suggest that most individuals recover within 3 months [29], [32], [37]. Autocorrelation was reduced using the median of EDSS scores within a quarter-year interval for individuals before model fitting.

The covariates in the model include time since onset, baseline characteristics, and DMT, including baseline covariates and time since onset interactions. Model comparisons suggests that accounting for CLOV provided a better fit for the data (Supplementary Table 2). However, due to outlying observations in time, the model did not converge for more complex models with many covariates, possibly because the level 1 variance became negative for outlying time observations. Moreover, variance of the residual was nonconstant (Supplementary Figure 2a), hence, time was truncated at 30 years to remove outlying observations. The AIC for the model with indicates that observational-level variance is a function of time, confirming that EDSS scores have higher variation at lower EDSS scale [38].

Progression in males was faster even though they had lower EDSS scores at onset (time to EDSS 6 was 20 years versus 25 years for females) in line with previous studies [39]–[43]. Individuals with below average age of onset took longer to reach EDSS milestones than individuals with above average age of onset, expected time to EDSS 6 was over 30 years versus 17 years, respectively. However, this does not imply more favourable outcome since younger individuals could still accumulate more disabilities at younger age [44], [45]. Individuals diagnosed historically progressed faster than those diagnosed more recently (time to EDSS 6 was 25 years and over 30 years for individuals diagnosed between 2007 – 2012, and >2012 – 2018, respectively) (Figure 2). This apparent improvement in outcome coincides with the era of high efficacy DMTs.

The combined effect of age of onset and sex when they were applied at 15 years post onset indicated faster progression in older individuals and males. Furthermore, the combined effects of year of diagnosis show that patients diagnosed historically progressed faster than patients diagnosed in 2007-2011, >2011 (Table 2).

In our cohort, the effect of DMT is uncertain because its effects depend on the model used (Supplementary Table 4). We attribute this effect to several sources of bias. Firstly, people with more aggressive MS are more likely to be selected for DMT. Secondly, people receiving DMT are often follow up more frequently; this was evident in our cohort where PwMS who had never received DMT had half as many EDSS measurements than those who received DMT. PwMS who did not receive DMT may therefore have reached key disability milestones years before their EDSS scores were measured. Thirdly, there may be many confounders between DMT and progression for which we have not adjusted.

The limitations of the study includes potential measurement bias which may be present in the EDSS scores, especially because of inter-rater variability is higher at lower scores [18]. This could affect the true disability score and trajectory of individuals. We did not have the ability to incorporate comorbidities such as depression and smoking, which could interact with MS outcome [46]–[50]. Due to the problem of extrapolation, we have compared expected EDSS at 10 years with year of diagnosis because the group of MS patients diagnosed after 2011 doesn’t have a follow up data of 15 years post diagnosis. There may be confounders of the relationships between age, sex, year of diagnosis and progression which we have not attempted to adjust for. We have not adjusted for covariates that may act as potential confounders for age, sex, year of diagnosis, and disease progression.

Compared to the earlier model [12], both models confirm that the progression of PwMS is nonlinear, even in a much larger contemporary sample. However, the current model adjusts for baseline covariates, and interactions. We found that males, older patients, and historical patients progress faster.

The modelling approach could be applied to other cohorts to estimate EDSS scores at certain time points after adjusting for baseline covariates. The ability to produce long term estimations gives this model great potential as an outcome measure to identify meaningful biomedical, genetic, and MRI biomarkers for clinical studies. For instance, estimated EDSS scores at latter years can be used as a phenotype in a genome-wide association studies (GWAS) of MS progression.

## Statement of Ethics

People with MS were selected from a regional disease registry according to accepted diagnostic criteria and were under the care of regional specialists in an MS centre in Cardiff, UK. A minority from neighbouring health boards (Aneurin Bevan, Swansea Bay, Powys) who had received some care or were previously resident in Cardiff were also included. Patients in the registry with a diagnosis of MS, a recorded date of MS onset, and who had consented to research were included.

## Conflict of Interest Statement

KEH reports speaker and personal fees from Roche, Merck, and Biogen, and travel grants to attend educational meetings from Roche, Novartis, Merck and Biogen.

ECT has received honoraria for consulting work from Novartis, Merck, Biogen and Roche, and funding to attend or speak at educational meetings from Biogen, Janssen, Merck, Roche, Takeda and Novartis.

NPR reports honoraria from Roche, Sanofi Genzyme and Novartis, and research grants from Novartis, Sanofi Genzyme and Biogen.

ML received fees for advising on a secondary analysis of an RCT sponsored by North Bristol NHS trust.

ECU declares no financial interests.

PH is a member of the Scientific Review Committee of Enroll-HD, for which he receives a small Honorarium.

## Funding Sources

ECU received salary from Cardiff University Wellcome Institution Strategic Funding award, Award Number: 204824/Z/16/Z. ML employed by University of Bristol, core-funded. PH is supported by the Medical Research Council (MRC) Centre for Neuropsychiatric Genetics and Genomics, Award Number: MR/K013041/1.

## Author contributions

PH, LJ, NPR and ECT contributed to conceptualization of the project; ECT, NPR, JH, ML, LJ, AND PH contributed to funding acquisition; ECU, KEH and ECT contributed to Data curation; ECU, ML, PH, and KLK contributed to methodology and formal analysis; ML, PH, and ECT contributed to supervision; ECU and ECT contributed to original draft; KEH, JH, KLK, PH, NPR, and ML contributed to reviewing & editing.

## Data Availability Statement

The dataset used in this study may be available upon request. Request should be made to the corresponding author.

## Notes

### Competing Interest Statement

The authors have declared no competing interest.

### Funding Statement

ECU received salary from Cardiff University Wellcome Institution Strategic Funding award. ML employed by University of Bristol, core-funded. PH is supported by the Medical Research Council (MRC) Centre for Neuropsychiatric Genetics and Genomics, Award Number

### Author Declarations

This study has been approved by the Research Ethics Committee of the School of Medicine, Cardiff University, Cardiff, United Kingdom.

